# *LRRK2* p.M1646T is associated with glucocerebrosidase activity and with Parkinson’s disease

**DOI:** 10.1101/2020.09.23.20197558

**Authors:** Yuri L. Sosero, Eric Yu, Lynne Krohn, Uladzislau Rudakou, Kheireddin Mufti, Jennifer A. Ruskey, Farnaz Asayesh, Sandra B. Laurent, Dan Spiegelman, Stanley Fahn, Cheryl Waters, S. Pablo Sardi, International Parkinson Disease Genomics Consortium (IPDGC), Sara Bandres-Ciga, Roy N. Alcalay, Ziv Gan-Or, Konstantin Senkevich

## Abstract

**Background and objectives:** The *LRRK2* p.G2019S Parkinson’s disease (PD) variant is associated with elevated glucocerebrosidase (GCase) activity in peripheral blood. We aimed to evaluate the association of other *LRRK2* variants with PD and its association with GCase activity.

**Methods:** *LRRK2* and *GBA* were fully sequenced in 1,123 PD patients and 576 controls from the Columbia and PPMI cohorts, in which GCase activity was measured in dried blood spots by liquid chromatography-tandem mass spectrometry.

**Results:** *LRRK2* p.M1646T was associated with increased GCase activity in the Columbia University cohort (β=1.58, *p*=0.0003), and increased but not significantly in the PPMI cohort (β=0.29, *p=*0.58). p.M1646T was associated with PD (OR=1.18, 95%CI=1.09-1.28, *p*=7.33E-05) in 56,306 PD patients and proxy-cases, and 1.4 million controls.

**Conclusions:** Our results suggest that the p.M1646T variant is associated with risk of PD with a small effect and with increased GCase activity in peripheral blood.

## Introduction

Parkinson’s disease (PD) is mostly caused by an interaction between genetic and environmental factors.^1^ Variants in *GBA* and *LRRK2* are among the most common genetic risk factors of PD.^2, 3^ The frequency of these variants varies in different populations, with *GBA* variants found in 5-20%^2^ and *LRRK2* variants reported in 1-40% of PD patients.^4^

The activity of the enzyme encoded by *GBA*, β-glucocerebrosidase (GCase), is reduced in carriers of *GBA* variants, but also in a subset of PD patients without *GBA* variants.^5, 6^ There are contradicting results regarding the effect of the *LRRK2* p.G2019S variant on GCase activity. In peripheral blood, this variant was associated with an increased activity,^6^ whereas in patient-derived dopaminergic neurons with *LRRK2* variants, GCase activity was reduced.^7^ A variant in *TMEM175*, p.T393M, has been associated with reduced GCase activity and, together with *GBA* variants and the *LRRK2* p.G2019S variant, explain only 23% of the variance in GCase activity in peripheral blood.^8^ These observations suggest that other genetic or environmental factors affect GCase activity.

In the current study, we performed full sequencing of *LRRK2* and *GBA* and examined the effect of common *LRRK2* variants on GCase activity in peripheral blood in two cohorts: from Columbia University and from the Parkinson’s Progression Markers Initiative (PPMI). We further examined the association of *LRRK2* variants identified through this analysis with risk of PD using data from the International Parkinson’s Disease Genomics Consortium (IPDGC), UK biobank and 23andMe, Inc. genome-wide association study (GWAS) meta-analysis.^9^

## Materials and methods

### Study population

To analyze the effects of *LRRK2* variants on GCase activity, two cohorts were included: 1) The Columbia University cohort (n=1,229, PD=797, Controls=432) and 2) The PPMI cohort (n=470, PD=326, Controls=144). Both cohorts have been previously described,^10, 11^ and their demographic data is detailed in Table 1. The Columbia cohort consisted of patients and controls of mixed ethnicity (mainly of European origin, including 308 individuals of Ashkenazi Jewish descent). Data on the effect of the *LRRK2* p.M1646T variant on risk of PD was extracted from the recent PD GWAS, including 37,688 PD patients, 18,618 UK Biobank proxy-cases and 1.4 million controls.^9^ All PD patients were diagnosed by movement disorder specialists according to the UK brain bank criteria^12^ or the MDS clinical diagnostic criteria.^13^

**Table 1.**
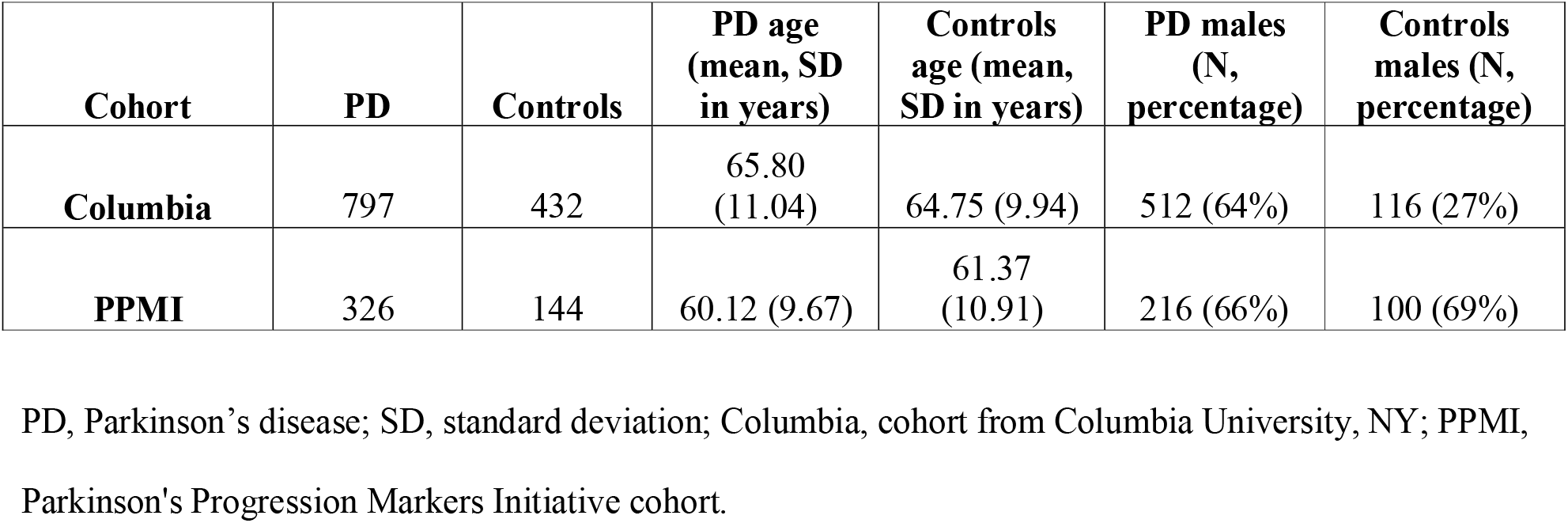
Demographic data of the cohorts to study *LRRK2* effect on GCase activity.

### Standard Protocol Approvals, Registrations, and Patient Consents

The institutional review boards approved the study protocols, and informed consent was obtained from all participants before entering the study. 23andMe participants provided informed consent and participated in the research online, under a protocol approved by the external AAHRPP-accredited IRB, Ethical & Independent Review Services (E&I Review).

### Genetic analysis

#### LRRK2 and GBA Sequencing in the Columbia University cohort

We performed full sequencing of *LRRK2* and *GBA* in the Columbia University cohort using targeted sequencing with Molecular Inversion Probes (MIPs) and Sanger sequencing as previously described.^14-16^ The full protocol and the library of MIPs used for sequencing *LRRK2* and *GBA* are available online (https://github.com/gan-orlab/MIP_protocol). A standard quality control protocol was performed as previously described,^17^ and the code is available at https://github.com/gan-orlab/MIPVar/.

#### Genetic data from PPMI and IPDGC

Due to the alignment difficulties with *GBA*, data on *GBA* variants in the PPMI cohort were extracted from combined data including whole genome sequencing data, whole exome sequencing data and RNA-seq as previously reported.^11^ Data on *LRRK2* p.G2019S, p.M1646T, p.N551K-p.R1398H and p.N2081D were extracted from imputed GWAS data (Illumina Immunochip and NeuroX arrays) downloaded from the PPMI project website (https://ida.loni.usc.edu/). To examine the association of *LRRK2* variants with PD, we extracted data from the recent PD GWAS meta-analysis.^9^

### GCase activity

Dried blood spots (DBS) were obtained as previously described.^18, 19^ GCase activity was measured in participants from Columbia University at Sanofi laboratories by liquid chromatography-tandem mass spectrometry (LC-MS/MS) from dried blood spots, as a part of multiplex assay with four additional lysosomal enzymes as previously described.^6, 20^ PPMI study participants donated blood on the first visit (baseline) and every year, which was frozen and stored in -80C freezer. Samples from the first three years of the cohort were thawed, and DBS were obtained. Activity was measured as previously described, using the mean GCase activity for each participant across all visits.^6, 11^

### Statistical Analysis

Linear regression models were used to test for association between *LRRK2* variants and GCase activity in the Columbia and PPMI cohorts, adjusting for age, sex, PD status, *GBA* status and ethnicity. In the PPMI cohort additional adjustment for white blood cells count was performed as suggested previously.^11^ We then repeated the analysis after excluding *LRRK2* p.G2019S and *GBA* variants carriers in both Columbia and PPMI cohorts. In addition, to examine whether there are sex-specific effects, we performed additional analyses stratifying the cohorts by sex (code available at https://github.com/gan-orlab/LRRK2_GCase). Bonferroni correction for multiple comparisons was applied as needed. All statistical analyses were performed using R or PLINK version 1.9.^21, 22^

### Results

In the Columbia University cohort, we identified 26 *LRRK2* common variants with MAF >1% (Supplementary Table 1), including 9 nonsynonymous variants, 12 intronic variants and 5 synonymous variants. We did not analyze rare variants, as the small number of carriers of these variants would not allow estimating their impact on GCase activity.

The *LRRK2* p.M1646T variant was associated with increased GCase activity compared to non-carriers (12.65 mmol/l/h vs. 11.38 mmol/l/h, respectively, β=1.58, *p=*0.0003, Table 2, Supplementary Table 1) in the Columbia University cohort. The effect of p.M1646T on GCase activity was stronger in PD (GCase=13.08 mmol/l/h, β=1.74, *p=*0.0011) and did not reach statistical significance in controls (GCase=11.96 mmol/l/h, β=1.37, *p=*0.068, Table 2, Supplementary Table 2-3). After exclusion of p.G2019S carriers, the association of p.M1646T with increased activity remained strong (GCase=12.64 mmol/l/h, β=1.73, *p=*6.24E-05, Supplementary Table 4-5). The *LRRK2* p.G2019S variant was associated with increased GCase activity as previously described.^6^ Two variants from the protective haplotype p.N551K-p.R1398H-p.K1423K were nominally associated with GCase activity, but this association was not statistically significant after Bonferroni correction (Table 2). We then demonstrated that the *LRRK2* p.M1646T variant was strongly associated with PD, using data from the recent PD GWAS meta-analysis,^9^ including 37,688 PD patients, 18,618 UK Biobank proxy-cases and 1.4 million controls (OR=1.18 95% CI=1.09-1.28, *p*=7.33E-05).

**Table 2.** Impact of *LRRK2* variants on GCase activity.

| *LRRK2* variants | N of carriers | Estimate | SE | p-value* | GCase_mean | Gcase_SD |
| --- | --- | --- | --- | --- | --- | --- |
| **Columbia cohort** |  |  |  |  |  |  |
| **PD + controls (N=1229)** |  |  |  |  |  |  |
| p.R1398H | 204 | 0.521 | 0.233 | 0.026 | 11.960 | 3.804 |
| p.M1646T | 58 | 1.578 | 0.431 | 0.0003 | 12.652 | 4.529 |
| p.G2019S | 61 | 1.370 | 0.438 | 0.0018 | 12.798 | 4.340 |
| **PD (N=797)** |  |  |  |  |  |  |
| p.R1398H | 123 | 0.495 | 0.298 | 0.097 | 11.776 | 3.863 |
| p.M1646T | 36 | 1.736 | 0.528 | 0.0011 | 13.078 | 4.600 |
| p.G2019S | 57 | 1.440 | 0.450 | 0.0014 | 12.877 | 4.471 |
| **Controls (N=432)** |  |  |  |  |  |  |
| p.R1398H | 81 | 0.755 | 0.383 | 0.050 | 12.240 | 3.720 |
| p.M1646T | 22 | 1.367 | 0.746 | 0.068 | 11.956 | 4.425 |
| p.G2019S | 4 | -0.030 | 1.719 | 0.986 | 11.658 | 1.286 |
| **PPMI cohort** |  |  |  |  |  |  |
| **PD + controls (N=470)** |  |  |  |  |  |  |
| p.R1398H | 61 | -0.724 | 0.340 | 0.034 | 10.936 | 2.961 |
| p.M1646T | 23 | 0.295 | 0.543 | 0.587 | 12.717 | 3.510 |
| p.G2019S | 6 | 0.004 | 1.050 | 0.997 | 11.453 | 2.648 |
| **PD (N=326)** |  |  |  |  |  |  |
| p.R1398H | 41 | -0.893 | 0.406 | 0.028 | 10.445 | 3.045 |
| p.M1646T | 17 | 0.073 | 0.623 | 0.907 | 12.385 | 3.259 |
| p.G2019S | 6 | -0.078 | 1.037 | 0.940 | 11.453 | 2.648 |
| **Controls (N=144)** |  |  |  |  |  |  |
| p.R1398H | 20 | -0.240 | 0.622 | 0.701 | 11.941 | 2.564 |
| p.M1646T | 6 | 1.169 | 1.082 | 0.282 | 13.657 | 4.333 |

SE, standard error; N, number; GCase\_mean, mean glucocerebrosidase activity, $\mu\text{mol/l/h}$ ; SD, standard deviation; PD, Parkinson's disease; Columbia, cohort from Columbia University, NY; PPMI, Parkinson's Progression Markers Initiative cohort; \*, Bonferroni correction significance threshold for Columbia cohort ( $\alpha=0.05/26=0.0019$ ) and ( $\alpha=0.05/5=0.01$ ) for PPMI cohort.

As a replication for GCase activity, we used data from the PPMI cohort, and analyzed the association of p.R1398H (representing the protective haplotype), p.M1646T, p.G2019S and p.N2081D with GCase activity (Table 2, Supplementary Table 8). The p.M1646T variant showed the same direction of effect and similar average GCase activity value as observed in the Columbia cohort, but did not reach statistical significance, possibly due to the small number of carriers (n=23), compared to non-carriers (12.72 mmol/l/h vs. 11.84 mmol/l/h, respectively, β=0.29, *p=*0.59; Table 2). Only six carriers of the *LRRK2* p.G2019S variant were included in the PPMI cohort, and the association of this variant with GCase activity, as well as of the protective haplotype, were not statistically significant after Bonferroni correction (Table 2, Supplementary Table 8). Stratified analysis by sex did not identify sex differences in GCase activity in both cohorts (Supplementary Table 6-8).

## Discussion

In the current study, we show that the *LRRK2* p.M1646T variant is associated with PD and with increased GCase activity in peripheral blood. The association of this variant with PD has been previously demonstrated,^3, 23^ and we now confirmed it in a larger European cohort. Despite its smaller effect on PD risk compared to the *LRRK2* p.G2019S variant, the effect of p.M1646T on GCase activity was larger than the effect of p.G2019S. However, since the results on GCase activity did not fully replicate in the PPMI cohort, additional studies are required to understand the associations between *LRRK2* variants, GCase activity and PD risk.

In a recent study, the *LRRK2* pathogenic variants p.G2019S, p.R1441G, and p.R1441C were associated with reduced GCase activity in patient-derived dopaminergic neurons, and correction of these variants resulted in normalization of GCase activity.^24^ Conversely, in the current study, deleterious *LRRK2* variants (p.G2019S and p.M1646T) were associated with increased GCase activity in peripheral blood. There are several potential explanations for these differences in the direction of effects on GCase activity, including: a) different effects of *LRRK2* variants in the central nervous system vs. peripheral blood, b) the possibility that iPSC-derived dopaminergic neurons, which are young cells, are different than patient tissues, due to the natural aging process, and c) the different methods used to measure GCase activity.

Considering the study suggesting that *LRRK2* variants are associated with reduced GCase activity,^24^ drugs targeting *LRRK2* activity could be repurposed for *GBA*-PD, and drugs that target GCase activity could be used for *LRRK2*-PD. However, this potential association between *LRRK2, GBA* and GCase activity should be carefully studied further, since other data suggests that *LRRK2* variants are not associated with reduced GCase activity. Patients with *GBA*-PD (and thus, reduced GCase activity) have a more severe phenotype with faster disease progression and cognitive decline, depression and anxiety, compared to sporadic PD.^25-27^ In contrast, *LRRK2* variants carriers have a milder phenotype with slower disease progression and lower frequency of cognitive symptoms compared to sporadic PD.^28, 29^ Moreover, two independent studies demonstrated that carriers of both *LRRK2* and *GBA* variants seem to have a benign phenotype, similar to those who carry *LRRK2* variants only.^30, 31^ If indeed *LRRK2* variants lead to reduced GCase activity as suggested,^24^ we would expect that patients with both *LRRK2* and *GBA* variants would have a severe phenotype. Instead, their phenotype is milder,^30, 31^ which may raise the hypothesis that the increased GCase activity we observed in peripheral blood may have some protective effect on PD phenotype. This hypothesis requires additional studies in human cohorts and disease models.

Our study has several limitations. In our cohorts, difference in sex between PD patients and controls was significant. To address this limitation, we adjusted the regression model with sex as covariate, as well as other covariates. The Columbia cohort included individuals of mixed ethnicity, mainly of European and Ashkenazi Jewish ancestry, and we adjusted the regression models for ethnicity. Another limitation is that GCase activity was measured in blood, which does not necessarily reflect GCase activity in the brain. More specifically, in the PPMI cohort it was measured from frozen blood, and the total number of carriers of *LRRK2* variants in this cohort was relatively low.

To conclude, we demonstrated that the *LRRK2* p.M1646T variant is associated with increased GCase activity in peripheral blood and with increased risk of PD. The interplay between *LRRK2, GBA* and GCase activity should be studied in additional cohorts and relevant disease models.

## Data Availability

Code available at https://github.com/gan-orlab/LRRK2_GCase.
Part of the data used in the preparation of this article were obtained from the Parkinson’s Progression Markers Initiative (PPMI) database (www.ppmiinfo.org/data). For up-to-date information on the study, visit www.ppmiinfo.org. PPMI a public private partnership is funded by the Michael J. Fox Foundation for Parkinson’s Research and funding partners, including (list the full names of all of the PPMI funding partners found at www.ppmiinfo.org/fundingpartners). We would like to thank the 23andMe research team. We would like to also thank all members of the International Parkinson Disease Genomics Consortium (IPDGC). For a complete overview of members, acknowledgements and funding, please see http://pdgenetics.org/partners.

## Acknowledgements

Data used in the preparation of this article were obtained from the Parkinson’s Progression Markers Initiative (PPMI) database (www.ppmiinfo.org/data). For up-to-date information on the study, visit www.ppmiinfo.org.““PPMI – a public-private partnership – is funded by the Michael J. Fox Foundation for Parkinson’s Research and funding partners, including (list the full names of all of the PPMI funding partners found at www.ppmiinfo.org/fundingpartners). We would also like to thank the research participants and employees of 23andMe for making this work possible. The full GWAS summary statistics for the 23andMe discovery data set will be made available through 23andMe to qualified researchers under an agreement with 23andMe that protects the privacy of the 23andMe participants. Please visit research.23andme.com/collaborate/ for more information and to apply to access the data. We would like to also thank all members of the International Parkinson Disease Genomics Consortium (IPDGC). For a complete overview of members, acknowledgements and funding, please see http://pdgenetics.org/partners. ZGO is supported by the Fonds de recherche du Québec - Santé (FRQS) Chercheurs-boursiers award, in collaboration with Parkinson Quebec, and by the Young Investigator Award by Parkinson Canada. The access to part of the participants for this research has been made possible thanks to the Quebec Parkinson’s Network (http://rpq-qpn.ca/en/). KS is supported by a post-doctoral fellowship from the Canada First Research Excellence Fund (CFREF), awarded to McGill University for the Healthy Brains for Healthy Lives initiative (HBHL). We thank Daniel Rochefort, Hélène Catoire, Clotilde Degroot and Vessela Zaharieva for their assistance.

## Authors’ Roles

1. Research project: A. Conception (YLS, ZGO, KS), B. Organization (YLS, EY, LK, UR, JAR, FA, KM, SBL, DS, SF, CW, SBC, RNA, ZGO, KS), C. Execution (YLS, EY, UR, LK, KM, JAR, FA, DS, KS).
2. Statistical Analysis: A. Design (YLS, ZGO, KS), B. Execution (YLS, KS).
3. Manuscript Preparation: A. Writing of the first draft (YLS, ZGO, KS), B. Review and Critique (YLS, EY, LK, UR, JAR, FA, KM, SBL, DS, SF, CW, SBC, PS, RNA, ZGO, KS).

## Financial Disclosures of all authors (for the preceding 12 months)

SF received consulting fees/honoraria for board membership from Retrophin Inc., Sun Pharma Advanced Research Co., LTD and Kashiv Pharma. CHW received research support from Sanofi, Biogen, Roche, consulting fees/honoraria from Amneal, Adamas, Impel, Kyowa, Mitsubishi, Neurocrine, US World Meds, Acadia, Acorda. RNA received consultation fees from Biogen, Denali, Genzyme/Sanofi and Roche. ZGO received consultancy fees from Lysosomal Therapeutics Inc. (LTI), Idorsia, Prevail Therapeutics, Inceptions Sciences (now Ventus), Ono Therapeutics, Denali and Deerfield. Rest of the authors have nothing to report.

